# COVID-19 mortality among working-age Americans in 46 states, by industry and occupation

**DOI:** 10.1101/2022.03.29.22273085

**Authors:** Yea-Hung Chen, Ruijia Chen, Marie-Laure Charpignon, Mathew V Kiang, Alicia R Riley, M Maria Glymour, Kirsten Bibbins-Domingo, Andrew C Stokes

## Abstract

**Introduction:** A small body of epidemiological research suggests that working in an essential sector is a risk factor for SARS-CoV-2 infection or subsequent disease or mortality. However, there is limited evidence to date on the US, or on how the risks associated with essential work differ across demographic subgroups defined by age, sex, and race/ethnicity.

**Methods:** Using publicly available data from the National Center for Health Statistics on deaths occurring in the US in 2020, we calculated per-capita COVID-19 mortality by industry and occupation. We additionally calculated per-capita COVID-19 mortality by essential industry—essential or not—by age group, sex, and race/ethnicity.

**Results:** Among non-military individuals and individuals with a known industry or occupation, there were 48,030 reported COVID-19 deaths, representing 25.1 COVID-19 deaths per 100,000 working-age individuals after age standardization. Per-capita age-standardized COVID-19 mortality was 1.96 times higher among essential workers than among workers in non-essential industries, representing an absolute difference of 14.9 per 100,000. Across industry, per-capita age-standardized COVID-19 mortality was highest in the following industries: accommodation and food services (45.4 per 100,000); transportation and warehousing (43.4); agriculture, forestry, fishing and hunting (42.3); mining (39.6); and construction (38.7).

**Discussion:** Given that SARS-CoV-2 is an airborne virus, we call for collaborative efforts to ensure that workplace settings are properly ventilated and that workers have access to effective masks. We also urge for paid sick leave, which can help increase vaccine access and minimize disease transmission.

## Introduction

A small body of epidemiological research suggests that working in an essential sector (a sector identified by local or regional government as being essential to critical functions, and thus exempt from stay-at-home orders) is a risk factor for SARS-CoV-2 infection or subsequent disease or mortality. An analysis of data from the UK Biobank project, for example, found that the risk of severe COVID-19 disease was higher among essential workers than among workers in non-essential sectors.^1^ Analyses of mortality data from the state of California show that essential workers have higher per-capita COVID-19 mortality, as compared to workers in non-essential sectors, particularly among those working in the agriculture, emergency, facilities, manufacturing, and transportation/logistics sectors. For example, between March 2020 and November 2021, annualized per-capita COVID-19 mortality among agriculture workers was 131.8 per 100,000, compared to 27.5 per 100,000 among non-essential workers.^2,3^ We hypothesize that these discrepancies are due, at least in part, to weaknesses in protections in workplace settings, particularly given documented instances of inadequate provision of personal protective equipment^4–6^ and poor ventilation.^7^ Incomplete coverage of paid sick leave^8,9^ and crowded housing^10^ may also heighten risk.

There is limited evidence to date on the US, or on how the risks associated with essential work differ across demographic subgroups defined by age, sex, and race/ethnicity. Analyses of occupational risk conducted in the UK suggest essential work is associated with especially high risk for COVID-19 mortality among all non-White race/ethnicity groups combined compared to White essential workers.^1^ However, the extent to which this finding generalizes to the US is currently unclear.

This study expands the geographical scope of the California analyses, examining COVID-19 mortality occurring in 2020 among working-age Americans in 46 states, by industry and occupation. We ask whether the disparities identified in the California analyses have occurred at the national level. We also examine per-capita COVID-19 mortality per industry by age, sex, and race/ethnicity, to examine whether disparities between essential workers and workers in non-essential industries differed across demographic subgroups.

## Methods

We obtained publicly available data from the National Center for Health Statistics on deaths occurring in the US in 2020. We restricted analysis to individuals 20–64 years of age (we used a lower bound of 20 rather than 18 to facilitate standardization). Data on industry and occupation were unavailable from 4 states: Arizona, Iowa, North Carolina, and Rhode Island; decedents from these states, and the District of Columbia and Puerto Rico, were dropped from analysis. We defined COVID-19 deaths using underlying cause of death.

We defined industry using two-digit National Health Interview Survey codes and defined occupation using four-digit US Census codes (both sets of codes were pre-existing in the data). In line with prior analyses of California data,^2,3^ which were guided by pandemic-era definitions, we grouped some industries as not essential to pandemic-era functions. In this analysis, the industries included in this category were: arts, entertainment, and recreation; education services; finance and insurance; information; management; other services; professional, scientific, and technical services; and real estate, rental, and leasing. In some analyses, we categorized industry into 2 groups: essential and not essential. Age groups were defined using five-year age increments. We measured race/ethnicity using a single-race variable and a Hispanic variable, focusing on the following groups: Asian or Pacific Islander, non-Hispanic Black, Hispanic, and non-Hispanic White.

We calculated age-standardized (using direct standardization to the WHO standard population) and unstandardized (“crude”) per-capita COVID-19 mortality by industry and occupation, using estimates of population size from the 2020 American Community Survey. We also examined differences in per-capita COVID-19 mortality between essential and non-essential workers by age, sex, and race/ethnicity.

We restricted our tables of per-capita COVID-19 mortality to individuals with listed non-military occupations due to concerns regarding whether military individuals are adequately captured in the American Community Survey. We additionally restricted our analysis of occupation to occupations with 100 or greater COVID-19 deaths, to focus on occupations with high total COVID-19 burden.

## Results

Among working-age individuals across the 46 states included in our analysis, there were 62,176 reported COVID-19 deaths in 2020. There were 261 reported COVID-19 deaths among military individuals and 13,885 reported COVID-19 deaths among individuals with an unknown/missing industry or occupation. Within the latter group, there were 4,465 reported COVID-19 deaths among individuals who had never worked or could not work (due to disability or being a patient or inmate); these represent 7.2% of all COVID-19 deaths considered in this analysis.

Among non-military individuals and individuals with a known industry or occupation, there were 48,030 reported COVID-19 deaths (Table 1), representing 25.1 COVID-19 deaths per 100,000 working-age individuals after age standardization (Table 1). Per-capita age-standardized COVID-19 mortality was 1.96 times higher among essential workers than among workers in non-essential industries, representing an absolute difference of 14.9 per 100,000. Across industry, per-capita age-standardized COVID-19 mortality was highest in the following industries: accommodation and food services (45.4 per 100,000); transportation and warehousing (43.4); agriculture, forestry, fishing and hunting (42.3); mining (39.6); and construction (38.7). Per-capita age-standardized COVID-19 mortality among agriculture workers was 2.72 times higher than among workers in non-essential industries, representing an absolute difference of 26.7 per 100,000.

**Table 1:**
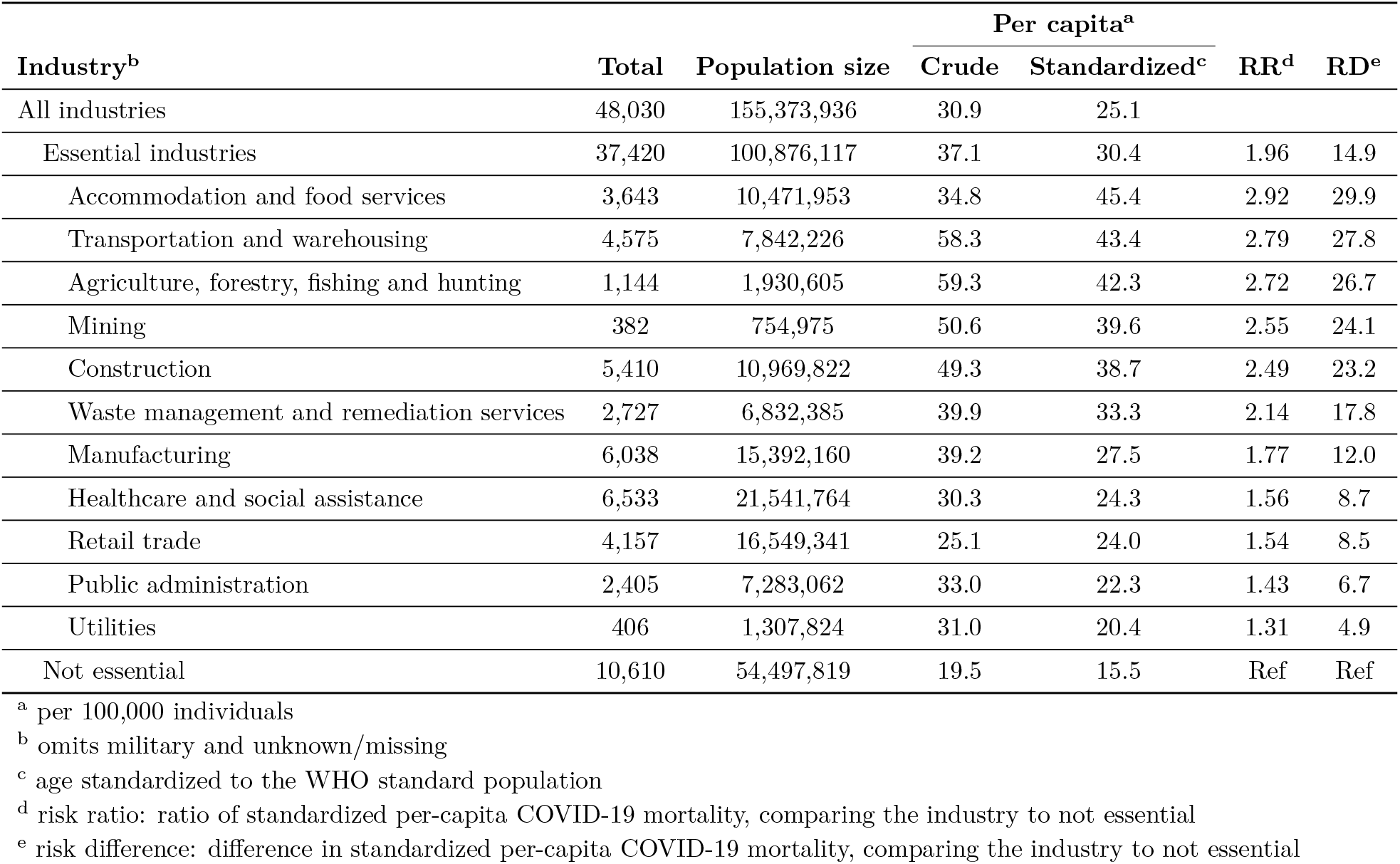
COVID-19 mortality working-age Americans (46 states), by industry, 2020.

**Table 2:** COVID-19 mortality among working-age Americans (46 states), by occupation, 2020.

| Occupation<sup>b</sup> | Total | Per capita<sup>a</sup> |  |  |
| --- | --- | --- | --- | --- |
|  |  | Population size | Crude | Standardized<sup>c</sup> |
| Construction laborers | 2,158 | 1,895,287 | 113.9 | 107.4 |
| Chefs and head cooks | 579 | 527,397 | 109.8 | 104.7 |
| Sewing-machine operators | 195 | 148,802 | 131.0 | 76.6 |
| Security guards and gaming surveillance officers | 781 | 966,190 | 80.8 | 75.9 |
| Roofers | 121 | 222,544 | 54.4 | 73.9 |
| Automotive service technicians and mechanics | 686 | 830,519 | 82.6 | 72.7 |
| Laborers and material movers | 1,770 | 2,541,210 | 69.7 | 71.8 |
| Clergy | 438 | 371,740 | 117.8 | 70.8 |
| Bailiffs, correctional officers, and jailers | 302 | 391,597 | 77.1 | 69.8 |
| Miscellaneous agricultural workers | 566 | 785,379 | 72.1 | 64.9 |
| Food processing workers, all other | 101 | 139,185 | 72.6 | 63.7 |
| Taxi drivers and chauffeurs | 509 | 566,412 | 89.9 | 63.6 |
| Bakers | 148 | 232,862 | 63.6 | 61.9 |
| Butchers and other meat-processing workers | 166 | 235,549 | 70.5 | 58.8 |
| Cooks | 1,185 | 2,165,739 | 54.7 | 56.4 |
| Musicians, singers, and related workers | 142 | 210,862 | 67.3 | 53.5 |
| Nursing, psychiatric, and home health aides | 1,314 | 2,060,128 | 63.8 | 52.7 |
| Production workers, all other | 905 | 1,365,611 | 66.3 | 51.6 |
| Grounds maintenance workers | 723 | 1,288,795 | 56.1 | 49.2 |
| Cleaners of vehicles and equipment | 156 | 374,033 | 41.7 | 48.9 |
| Dishwashers | 129 | 295,514 | 43.7 | 48.7 |
| Industrial truck and tractor operators | 378 | 690,313 | 54.8 | 48.0 |
| First-line supervisors of construction and extraction | 460 | 653,079 | 70.4 | 46.5 |
| Packaging and filling-machine operators and tenders | 160 | 276,399 | 57.9 | 46.0 |
| Driver/sales workers and truck drivers | 2,299 | 3,516,318 | 65.4 | 44.9 |

<sup>a</sup> per 100,000 individuals
<sup>b</sup> restricted to the 25 non-military occupations with the highest age-standardized per-capita COVID-19 mortality
<sup>c</sup> age standardized to the WHO standard population

By occupation, age-standardized per-capita COVID-mortality was highest among construction laborers (107.4 per 100,000); chefs and head cooks (104.7); sewing-machine operators (76.6); security guards and gaming surveillance officers (75.9); and roofers (73.9).

Occupations with the 25 highest number of COVID-19 deaths that were not among the 25 occupations with the highest age-standardized per-capita COVID-19 mortality include: janitors and building cleaners (1,426 COVID-19 deaths); retail salespersons (1,043); first-line supervisors of retail sales workers (900); registered nurses (862); K–12 teachers (808); all other managers (763); maids and housekeeping cleaners (660); personal care aides (659); carpenters (565); and customer service representatives (435).

Among both essential workers and non-essential workers, per-capita COVID-19 mortality increased with age (Figure 1). Across age groups, the ratio in per-capita COVID-19 mortality between essential and non-essential workers was greatest at ages 45–49 (2.18) while the difference between in per-capita COVID-19 mortality between essential and non-essential workers was greatest at ages 60–64 (69.3). Per-capita COVID-19 mortality was higher among essential workers than non-essential workers, regardless of age. This pattern held in analyses additionally stratified by sex and race/ethnicity (Figure 2). In these stratified analyses, differences in per-capita COVID-19 mortality between essential and non-essential workers were especially large among older workers and among non-Hispanic Black and Hispanic essential workers. These differences were observed for both males and females.

**Figure 1:**
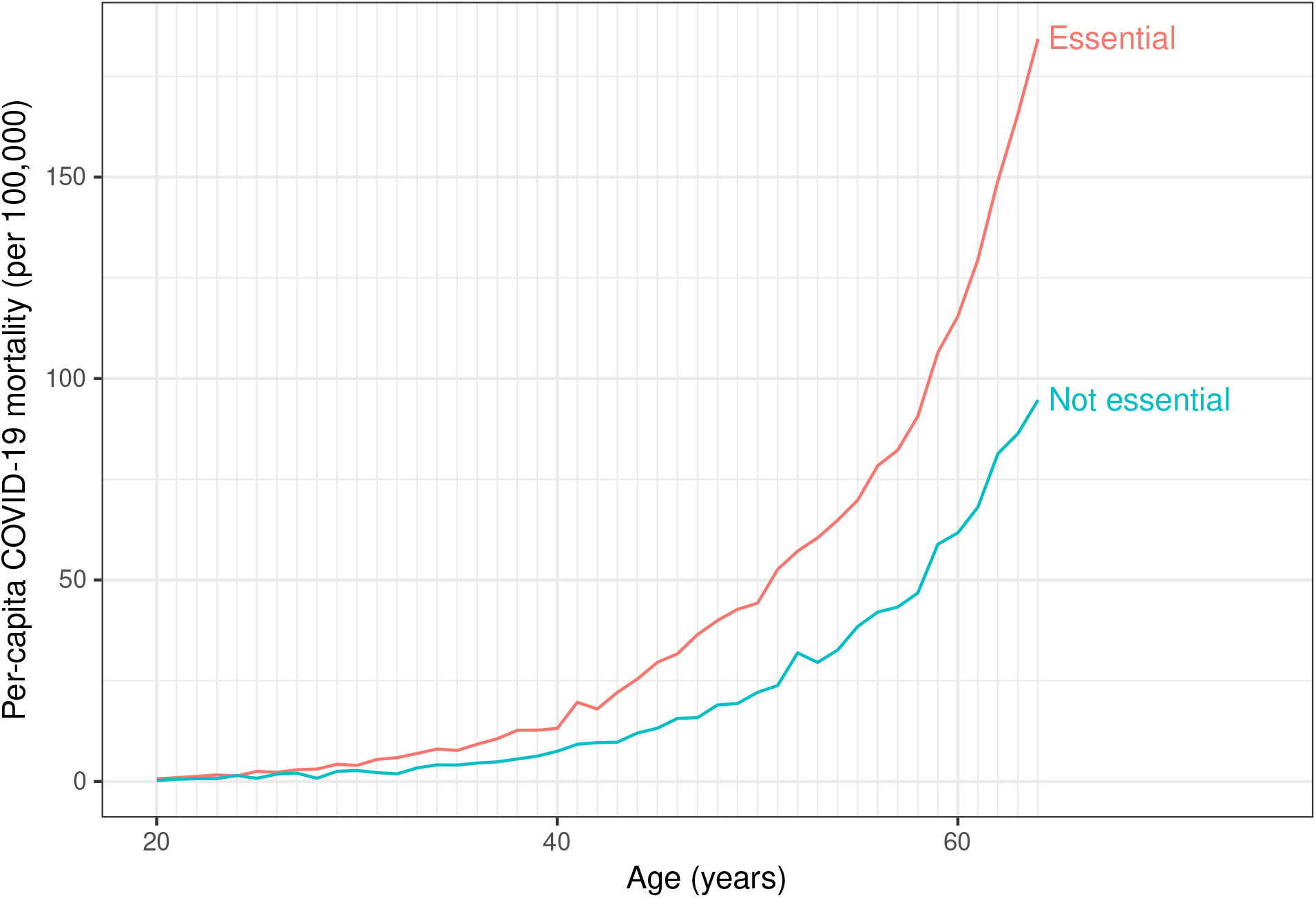
Per-capita COVID-19 mortality by industry (essential or not essential) and age, United States (46 states), 2020.

**Figure 2:**
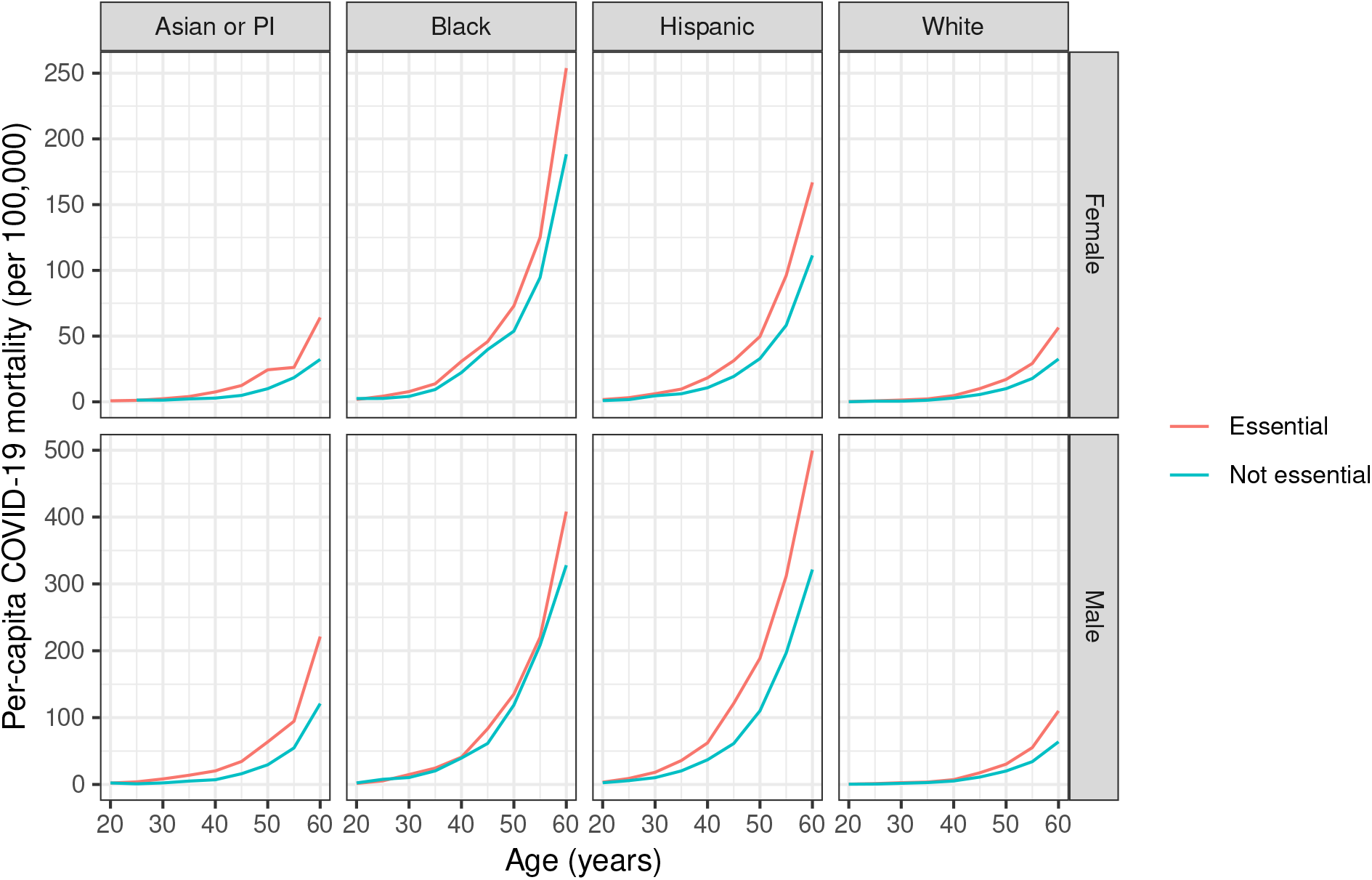
Per-capita COVID-19 mortality by industry (essential or not essential), age group, sex, and highest level of educational attainment, United States (46 states), 2020.

## Discussion

Among working-age Americans, per-capita COVID-19 mortality in 2020 was highest among essential workers, particularly those in the following industries essential industries; accommodation and food services; transportation and warehousing; agriculture, forestry, fishing and hunting; and mining industries. Essential workers had higher per-capita mortality than workers in non-essential industries, even after accounting for demographic variables. Differences in the disparity between essential workers and workers in non-essential industry across race/ethnicity groups might be attributable to heterogeneity in the geographies, industries, or occupations represented; structural factors such as crowded housing may also be relevant.

Our findings of disparities in COVID-19 mortality across occupational groups is consistent with earlier analysis of data from California,^2^ and indicate that the state was not unique in inadequately protecting essential workers. We acknowledge that the elevated risks among workers in essential industries may not be solely due to workplace transmission. However, we argue that work is irremovable from our interest in protecting essential workers and our mechanisms for doing so. Given that SARS-CoV-2 is an airborne virus,^11,12^ we call for collaborative efforts to ensure that workplace settings are properly ventilated and that workers have access to effective masks.^13,14^ We also urge for paid sick leave,^8^ which can help increase vaccine access and minimize disease transmission.

## Data Availability

Data are publicly available via the NCHS website.

https://www.cdc.gov/nchs/

## Notes

### Competing Interest Statement

The authors have declared no competing interest.

### Funding Statement

This study did not receive any funding

### Author Declarations

Only existing public datasets were used.

### Summary of Updates

Revisions to both Table 1 (adding public administration) and Table 2 (to standardized numbers)

